# Personalized, EEG-controlled intermittent theta burst stimulation

**DOI:** 10.64898/2026.04.27.26351877

**Authors:** Felix A. Maldonado Osorio, Mina Elhamiasl, Mary K. Gacek, Stewart A. Shankman, Ivan Alekseichuk

## Abstract

Brain-state-controlled transcranial magnetic stimulation (TMS) studies with real-time electroencephalography (EEG) show that the phase of ongoing oscillations modulates cortical susceptibility to TMS pulses. Translating this principle to repetitive clinical protocols, such as intermittent theta burst stimulation (iTBS), is an open challenge because within-train stimulation pulses corrupt real-time EEG. Moreover, the general difficulty of predicting EEG theta phase even to initiate an iTBS train applies. We present our solution for prefrontal EEG-phase-controlled iTBS, a personalized stimulation framework. We demonstrate the technical feasibility of aligning each train’s initial bursts to the individual prefrontal theta phase and propose a “seed-and-sustain” hypothesis, whereby intra-train stimulation-induced entrainment at the individual theta rhythm carries the later bursts. Future human trials will be needed to evaluate the practical benefits of this approach.

**Highlights:**

- Intermittent TBS synchronized to the prefrontal EEG theta rhythm is feasible
- Personalized iTBS-EEG parameters are stable over the typical session time
- Clinical evaluation of iTBS-EEG is a direction for future work

---

Brain state-controlled and closed-loop approaches to transcranial magnetic stimulation (TMS) have emerged as a promising frontier in precision psychiatry [1]. Multiple groups have demonstrated that the phase of ongoing electroencephalographic (EEG) oscillations modulates cortical susceptibility to a standard TMS pulse, with most evidence to date coming from recordings of evoked potentials in the motor cortex [2]. These findings raise the prospect that EEG-synchronized stimulation could reduce inter-individual variability in therapeutic neuromodulation. However, a methodological gap exists between modern EEG phase-controlled TMS paradigms and the repetitive stimulation protocols used clinically. The former deliver one or a few pulses timed to a target oscillatory phase to probe immediate effects [3], whereas the latter aim to induce plasticity with long stimulation trains. Such as, intermittent theta burst stimulation (iTBS) over the prefrontal cortex, used for depression treatment, delivers 10 bursts (triplets) at 5 Hz over a 2-second train with an 8-second inter-train interval [4]. Although the protocol’s burst rate was designed to mimic endogenous theta rhythms, synchronization of iTBS to ongoing EEG theta remains an open challenge. Two obstacles stand in the way. First, the rapid succession of stimulation pulses within each train corrupts EEG recordings, degrading real-time phase tracking after the first pulse. Second, prefrontal theta oscillations are less stationary and of lower amplitude than the motor alpha rhythms typically targeted in prior closed-loop work, demanding more robust [5] and computationally expensive methods. Overcoming these constraints would enable investigation of the clinical potential of state-controlled iTBS.

Here we present our method for EEG state-controlled iTBS (iTBS-EEG). We demonstrate its technical validity in the prefrontal cortex, enabled by a fast, parallelized system architecture and hierarchical optimization of individual state-control parameters. We also discuss the mechanistic rationale that underlies this method, which we term “seed-and-sustain.”

The core concept is to synchronize iTBS delivery to the individual theta oscillation, which originally inspired the protocol [4]. This achieves two goals: i. matching the inter-burst rate to the patient’s individual theta rhythm, and ii. delivering all pulses at the oscillatory phase most susceptible to perturbation. The former has shown promise in open-loop delivery [6]. The latter has preliminary support from a single-burst EEG-controlled stimulation study [7].

The method is described in Figure 1A-B and in supplementary materials. The main challenge is forecasting the biomarker (theta phase), which is needed because of technical delays, edge-of-data uncertainty, and correct burst alignment. Each burst must be triggered before the biomarker occurs, so that the burst’s mid-point coincides with it. The core logic is to first calibrate the algorithm on the individual EEG (i.e., stimulate virtually) and then apply calibrated parameters during the stimulation stage to forecast when to trigger a stimulation train. We keep the forecasting short and simple. The algorithm only forecasts the next occurrence of the biomarker based on the calibrated optimal anchor event (preceding theta peak or trough). It uses a calibrated inter-burst interval (IBI) to schedule subsequent bursts within the train. Our total system delay from EEG sampling to TMS coil discharge is 1.7-1.9 ms and processing speed is 0.1-0.4 ms per sample. We partly compensate for edge-of-data uncertainty by adaptive edge removal and calibrated forecasting based on ground truth during a calibration stage.

**Fig. 1.**
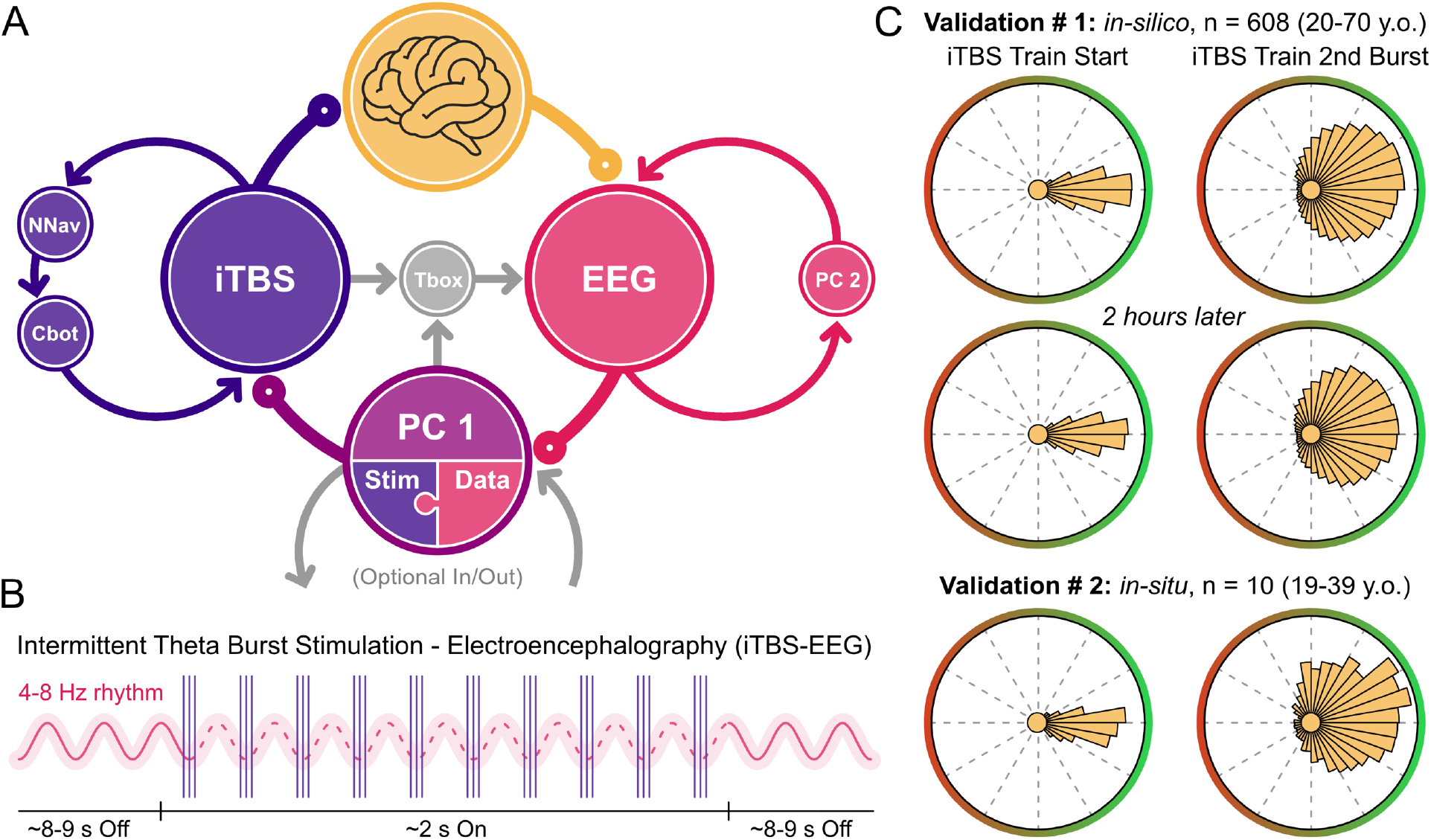
The personalized, state-controlled intermittent theta burst stimulation and electroencephalography (iTBS-EEG) framework (panel A) consists of 1. a high-dynamic range EEG unit with real-time data streaming, 2. a computer unit “PC 2” for EEG monitoring and governing, 3. a computer unit “PC 1” for state control, including parallel software processes for i. data processing and ii. stimulation pulse control, and 4. a TMS unit with 5. real-time neuronavigation and 6. a cobot for coil positioning. Optionally, the EEG unit can receive external event markers via a trigger box, and computer units can receive and send synchronization signals to a cognitive task computer. Panel B depicts protocol schematics, where theta-burst timings, inter-burst interval (IBI), and, by extension, stimulation train length (∼ 2 s) are personalized based on each participant’s dominant theta rhythm. The experiment starts with a brief EEG recording and a system calibration stage to estimate individual real-time parameters before proceeding to the stimulation stage. Panel C reports the stimulation accuracy for the EEG theta phase-controlled iTBS targeting the left prefrontal cortex across two experiments. The accuracy is shown on the median normalized polar plots as a deviation from the targeted phase, where 0 degrees corresponds to the best accuracy (bars point to the ‘east’).The left column depicts iTBS train initiation accuracy (which depends on theta phase forecasting) and the right column shows the 2nd burst in the train (which depends on the calibrated IBI). Experiment 1 is the *in-silico* validation in the Dortmund Vital Study dataset (n = 608 participants). The system was calibrated using pre-task resting-state recordings and validated against pre-task (upper row) or post-task recordings (2 hours later; middle row). Experiment 2 is the *in-situ* validation in our experimental group (n = 10 participants; lower row).

We validated iTBS-EEG in two experiments, targeting theta rhythm (4-8 Hz) phase in the left prefrontal cortex. Experiment 1 virtually simulated phase-controlled stimulation on the Dortmund Vital Study dataset (n = 608, 20-70 years old, [8]), which includes two 3-min resting-state recordings per person made 2 hours apart. We calibrated the algorithm on the first recordings and applied it to both. Here, we report the phasic (cosine) accuracy of virtually stimulating at the theta phase of choice. We separately report the accuracy of initiating the stimulation train and of targeting the next theta cycle with the 2nd stimulation burst in the train (Fig. 1C). For the train initiation, the median accuracy was 96.2% in the first recording per participant and 95.1% in the second recording. For the 2nd burst alignment, the accuracy was 70.2% and 69.0%, respectively. Thus, the algorithm is stable over two hours post-calibration and reliable at each train’s start, with no significant differences (all pairwise, uncorrected p-values > 0.2) across age (20-35 / 35-50 / 50-70 years old) or sex. Experiment 2 was conducted at Northwestern University (n = 10, 19-39 years old; IRB# STU00223602) with all equipment active but TMS pulses withheld, enabling reliable EEG phase estimation (as done before [5]). Results, reported immediately post-calibration, agreed with Experiment 1: 92.0% median accuracy for train initiation and 70.9% median accuracy for the 2nd burst. Notably, phase accuracy for train start reflects genuine alignment of the first burst to the endogenous theta oscillation. However, for subsequent bursts, the accuracy will likely also depend on the consistency of stimulation-induced entrainment (discussed below), which is outside the scope of the current technical validation.

A practical consideration is that no EEG-controlled method can reliably forecast theta phase for several seconds ahead to maintain alignment throughout an iTBS train. We argue that aligning the initial bursts is likely enough. Intracranial recordings show that repetitive stimulation bursts produce frequency-specific entrainment persisting briefly past offset [9]. Phase-aligning the first few bursts within the train should therefore seed the intra-train cascade at optimal excitability. The subsequent bursts are delivered at the person’s individual theta rhythm, thereby riding the individual dynamics sustained by entrainment from the preceding rhythmic bursts – a “seed-and-sustain” mechanism. Indeed, personalizing iTBS to the individual theta rhythm has been shown beneficial even in open-loop delivery [6]. Single-burst theta-phase-triggered TMS [7] and recent direct hippocampal stimulation [10] further demonstrate that theta-phase-locked stimulation enhances electrophysiological connectivity. Our proposed mechanism combines rationales of personalized rhythm, intra-train entrainment, and phase-locked delivery.

In summary, we show the technical feasibility of EEG state-controlled iTBS over the prefrontal cortex. By aligning each stimulation train’s initial bursts to the individual theta rhythm and hypothesizing that the entrainment sustains subsequent bursts, we sidestep the multi-second forecasting problem. Clinical evaluation of this approach is a natural next step.

## Data Availability

The data used in the present study are available online (for Experiment 1) or upon reasonable request to the authors (for Experiment 2).

https://openneuro.org/datasets/ds005385/versions/1.0.2

## CRediT authorship contribution statement

**Felix A. Maldonado Osorio:** Software, Validation, Writing - Review & Editing; **Mina Elhamiasl:** Validation, Investigation, Writing - Review & Editing; **Mary K. Gacek:** Resources, Writing - Review & Editing; **Stewart A. Shankman:** Writing - Review & Editing, Supervision, Project administration; **Ivan Alekseichuk:** Conceptualization, Methodology, Software, Validation, Writing - Original Draft, Visualization, Supervision, Project administration, Funding acquisition.

## References

[1] Zrenner C, Ziemann U. Closed-Loop Brain Stimulation. Biological Psychiatry 2024;95:545– 52. 10.1016/j.biopsych.2023.09.014.

[2] Wischnewski M, Shirinpour S, Alekseichuk I, Lapid MI, Nahas Z, Lim KO, et al. Real-time TMS-EEG for brain state-controlled research and precision treatment: a narrative review and guide. J Neural Eng 2024;21:061001. 10.1088/1741-2552/ad8a8e.

[3] Varone G, Biabani M, Tremblay S, Brown JC, Kallioniemi E, Rogasch NC. The golden age of online readout: EEG-informed TMS from manual probing to closed-loop neuromodulation. NeuroImage 2025;322:121543. 10.1016/j.neuroimage.2025.121543.

[4] Blumberger DM, Vila-Rodriguez F, Thorpe KE, Feffer K, Noda Y, Giacobbe P, et al. Effectiveness of theta burst versus high-frequency repetitive transcranial magnetic stimulation in patients with depression (THREE-D): a randomised non-inferiority trial. The Lancet 2018;391:1683–92. 10.1016/S0140-6736(18)30295-2.

[5] Shirinpour S, Alekseichuk I, Güth MR, Haigh Z, Wischnewski M, Opitz A. Bayesian Temporal Prediction: A Robust Algorithm for Real-time EEG Phase-dependent Brain Stimulation. IEEE Transactions on Biomedical Engineering 2025. 10.1109/TBME.2025.3589970.

[6] Chung SW, Sullivan CM, Rogasch NC, Hoy KE, Bailey NW, Cash RFH, et al. The effects of individualised intermittent theta burst stimulation in the prefrontal cortex: A TMS-EEG study. Human Brain Mapping 2019;40:608–27. 10.1002/hbm.24398.

[7] Gordon PC, Belardinelli P, Stenroos M, Ziemann U, Zrenner C. Prefrontal theta phase-dependent rTMS-induced plasticity of cortical and behavioral responses in human cortex. Brain Stimulation 2022:118159. 10.1016/j.brs.2022.02.006.

[8] Gajewski PD, Getzmann S, Bröde P, Burke M, Cadenas C, Capellino S, et al. Impact of Biological and Lifestyle Factors on Cognitive Aging and Work Ability in the Dortmund Vital Study: Protocol of an Interdisciplinary, Cross-sectional, and Longitudinal Study. JMIR Res Protoc 2022;11:e32352. 10.2196/32352.

[9] Solomon EA, Sperling MR, Sharan AD, Wanda PA, Levy DF, Lyalenko A, et al. Theta-burst stimulation entrains frequency-specific oscillatory responses. Brain Stimulation 2021;14:1271– 84. 10.1016/j.brs.2021.08.014.

[10] Kragel JE, Lurie SM, Issa NP, Haider HA, Wu S, Tao JX, et al. Closed-loop control of theta oscillations enhances human hippocampal network connectivity. Nat Commun 2025;16:4061. 10.1038/s41467-025-59417-7.

