## Supplementary Methods for "Personalized, EEG-controlled intermittent theta burst stimulation"

iTBS-EEG hardware. The system comprises an EEG amplifier, a control computer, and a TMS machine. Bracketed text throughout this supplement indicates our specific implementation choices [device models and settings]. The EEG amplifier should provide a high dynamic range and a high sampling rate [10 kHz] to capture non-harmonic noise from the TMS system and should support parallel data streaming to two computers [BrainProducts™ ActiChamp Plus with TurboLink and ActiCap 64 ch]. Dedicating a second computer solely to continuous EEG recording and amplifier configuration frees resources on the main control computer. The control computer should receive data in real time with minimal delay [UDP] and issue fast triggering commands to the TMS system [TTL]. The TMS system should support high-rate repetitive stimulation, noise filtering or recharge-delay control, and external triggering [MagVenture™ x100 MO]. Movement of the TMS coil relative to the EEG electrodes during delivery should be minimized: even minor shifts alter the spatial profile of induced noise and risk mechanically perturbing the electrodes, both of which degrade data quality and state-control accuracy. We use a neuronavigated collaborative robot [Axilium Robotics™ Cobot v2 with Localite™ Navigator Research Premium].

iTBS-EEG software. The control computer runs on Ubuntu™ 24.04 LTS with the real-time Linux™ kernel. All operations are performed in MATLAB™ 2025b. The raw EEG data are recorded at a 10 kHz sampling rate and downsampled to 1 kHz on the control computer for EEG state-controlled processing at a millisecond timescale. The algorithm operates on a sample level, and the total software time cost is 0.1-0.4 ms (min-max) per sample. The total system delay from the recorded EEG sample at the electrode level to control computer to TMS coil discharge is 1.7-1.9 ms, which is dominated by the hardware delay of EEG data streaming. This was measured 600 times during a 10-min experiment as the delay between the single-pulse TMS triggering command issued by the control computer and the TMS artifact in the real-time EEG data seen at the same control computer. The software and hardware delays are non-additive: while one data sample is processed by the control computer, the next sample is recorded and pushed into the control's computer buffer by the EEG sub-loop.

The user specifies the brain area of interest by selecting the EEG electrode of interest and its reference points [Laplacian montage surrounding the electrode of interest], which act as a first-order spatial filter. Further, the user provides with the frequency band of interest [4-8 Hz] and the target phase [peak or trough]. The user also provides the number of pulses per burst [3], bursts per train [10], and trains per treatment [20 trains = 600 pulses]. The session then proceeds in two stages: system calibration and active stimulation, described below as implemented in our setup.

*Calibration stage.* During calibration, a brief EEG recording (4 min) is made while the participant is in the "reference" state – that is, engaged in the same activity as during subsequent iTBS delivery [resting state]. The recording is duplicated into a ground-truth copy and an optimization copy, used for virtual simulation and parameter estimation. The ground-truth copy is first filtered with a forward-backward high-pass IIR filter [0.75 Hz, 4th order] and cleaned with Artifact Subspace Reconstruction at a permissive cutoff [SD threshold of 40], chosen to preserve all physiological signal while removing gross artifacts. In practice, the exact threshold in the range of 20-80 had no systematic impact on the validation results. Then, a selected passband of interest [4-8 Hz] is extracted using a zero-phase Kaiser-windowed FIR filter [stopband attenuation of 80 dB]. From the optimization copy, we estimate a voltage threshold on Fp1/Fp2 electrodes to detect ocular artifacts [90th percentile] and a power threshold at the target electrode below which the current data epoch will be rejected as having too weak oscillation to yield a reliable phase estimate [median power split based on the calibration recording]. The optimization copy is processed as EEG will be processed during the active stimulation stage. EEG data are epoched and filtered [4-8 Hz] using fast FFT-based brickwall filtering. We then simulate the iTBS-EEG pipeline against the ground-truth and use Bayesian optimization to identify the optimal epoch length, epoch edge-removal length, anchor event (the reference phase from which the next occurrence of the target phase is forecasted), and forecasting distance, with the objective of maximizing agreement between the phase stimulated by the first burst in each train and the target phase extracted from the ground truth. A second optimization stage then identifies the inter-burst interval (IBI) that best sustains phase alignment across the remainder of the train.

*Active stimulation stage.* During active stimulation, the parameters identified in calibration are used to deliver iTBS trains aligned to the EEG phase of interest. At the software level, on the main control computer, data processing and stimulation triggering (one trigger per pulse or per theta-burst) run as two parallel processes to make best use of computer resources. Both software processes receive the corresponding calibrated parameters and user-defined choices at their first initiation. The data process has access to the input port [Ethernet] only: it captures the EEG data, downsamples to 1 kHz, re-references to the selected montage, applies fast bandpass filtering [4-8 Hz], applies the calibrated thresholds, and identifies the next stimulation moment, which it issues as a fire command. The stimulation process has access to the output port [TTL] only. While in the standby state, it listens for a fire command from the data process; upon receipt, it transitions to the fire state, in which it is no longer reachable from the processing loop. In the fire state, it delivers the iTBS train according to the calibrated parameters (forecasting distance and IBI) and user-defined number of bursts per train. Then, the stimulation process enforces an inter-train interval of at least 8 seconds (per the iTBS specifications) and checks how many trains have already been delivered before returning to standby. During firing, the stimulation process uses robust "tic-toc" logic; meanwhile, the data process continues to capture EEG data without interruption.
